# EEG Based Oculographic Analysis of Epileptic Nystagmus

**DOI:** 10.1101/2023.10.02.23296289

**Authors:** Aybuke Acar, Anthony Zampino, Neel Fotedar

**Affiliations:** Epilepsy Center, Neurological Institute, University Hospitals Cleveland Medical Center, Cleveland, OH, USA 44106; Department of Neurology, Case Western Reserve University School of Medicine, Cleveland, OH, USA 44106

**Keywords:** epileptic nystagmus, EEG, time constant, parieto-occipital

## Abstract

Epileptic nystagmus (EN) is a subtle seizure semiology, most commonly seen in seizures originating in the posterior cortical regions. EN is broadly categorized into type I and type II. Type I EN consists of contralateral repetitive saccadic eye movements alternating with post-saccadic slow drifts with an overall contralateral deviation. Type II EN is characterized by ipsilateral slow drift alternating with contralateral corrective saccades. In this article, we report a method to perform oculographic analysis of eye movements using EEG only. We used this method to classify the type of EN in three patients with parieto-occipital seizures.

In all three patients, the ictal EEG demonstrated repetitive saccadic eye movements, directed contralateral to the seizure onset zone. With prolonged time constant, we were able to identify this eye movement pattern as EN with distinct slow and fast phases. We were able to further characterize the type of EN as type I and type II. In all three patients, the direction of EN (direction of fast phase or saccades) was contralateral to the seizure onset zone.

EN can be easily missed on video-electroencephalography (vEEG) recordings because of various reasons. Our study demonstrates a systematic method of eye movement analysis on EEG, which can be used to not only identify EN as seizure semiology but also classify it, without requiring additional electrodes.

## 1. Introduction

Epileptic nystagmus (EN) is a rarely reported seizure semiology, most commonly associated with seizures originating in the posterior cortical regions (Kaplan and Tusa, 1993). It is characterized by fast (saccadic) repetitive eye movements, alternating with slow movements, with the fast phase being directed away from the seizure onset zone (Tusa et al., 1990).

EN is mainly classified into two types depending upon the underlying mechanism. Type I EN is postulated to originate from the activation of the cortical areas responsible for saccadic eye movements producing contraversive saccades. Type II EN is generated by the activation of cortical areas responsible for smooth pursuit (temporo-parieto-occipital junction), resulting in an ipsiversive slow phase alternating with a resetting contraversive saccade (Kaplan and Tusa, 1993; Tusa et al.,1990).

The eye acts like an electrical dipole with the anterior being more positive and the posterior being more negative. This creates a characteristic artifact on EEG depending on the type and direction of the eye movement (Lüders and Noachtar, 2000). For example, a leftward eye movement will create a large positivity at F7 and a simultaneous negativity at F8. In addition, the saccadic eye movements have a characteristic morphology with a rapid rise of potential, followed by a slow return to baseline, which is determined by the time constant (TC) (Lüders and Noachtar, 2000).

In this article, we aim to report a systematic method to analyze eye movement artifact on EEG, which could help identify EN and distinguish between its different types, without requiring additional electro-oculogram (EOG) electrodes.

## 2. Methods

We identified three patients from our database with parietal, occipital or parieto-occipital seizures and reported semiology of epileptic nystagmus, who had at least 24 hours of vEEG data available for review, using the Neurofax EEG-1200 system by Nihon Kohden (Tokyo, Japan). Eyes were not visible in the video in any of these patients. The report of EN was based on either bedside examination or from history.

First, we analyzed the EEG epoch of interest with the conventional settings of a TC of 0.1 second and a high-frequency filter of 70Hz in a longitudinal bipolar montage. We then analyzed 30 seconds of pre-ictal EEG for any unusual eye movements, followed by the ictal EEG. In the ictal EEG, we identified epochs of unilaterally directed repetitive saccadic eye movements. Following this, we analyzed the same epochs with a prolonged TC of ≥2 seconds to identify and characterize any slow eye movements, alternating with the saccadic eye movements.

In addition, we also characterized a single-channel oculographic waveform using the same settings of prolonged TC. By connecting F7 to an electrode relatively indifferent to horizontal eye movements (e.g. Cz or Fp1), we were able to study the characteristics of the slow phase and the fast phase.

Slow phase waveform was characterized as linear or non-linear, depending upon the slope of the waveform as it would approach the EEG baseline. A negative exponential decay of the slope of the waveform as it approaches the baseline is consistent with decreasing velocity as the eyes approach the midline. In addition, we also identified whether the slow and fast eye movements crossed the midline or not by analyzing whether the respective waveforms would cross the EEG baseline or not. For example, if the eyes start at midline and then drift toward the left by 20 degrees, the EEG would show a certain positive voltage at F7 away from the baseline. If the eyes then come back to the right by the same amount, the waveform would simply return to the baseline but it would not cross it. If instead the eyes would cross the midline and actually turn to the right side by another 20 degrees, the corresponding EEG waveform would be twice the amplitude of the initial waveform, hence crossing the EEG baseline. This analysis is only possible with the settings of prolonged TC.

This study was approved by the Institutional Review Board of University Hospitals Cleveland Medical Center.

## 3. Results

Patient #1 and #3 were monitored for five and three days, respectively, and only one seizure was captured for each of them during the recording. Patient #2 was monitored for one week. During the initial five days, he had very frequent seizures at ∼9-13/hour. All of these seizures had the EEG characteristics consistent with nystagmus, as described below.

Patient demographics, nystagmus characteristics and EEG findings are summarized in table 1. In all three patients, there was no specific pattern of repetitive eye movement artifact in the immediate pre-ictal period.

**Table 1:**
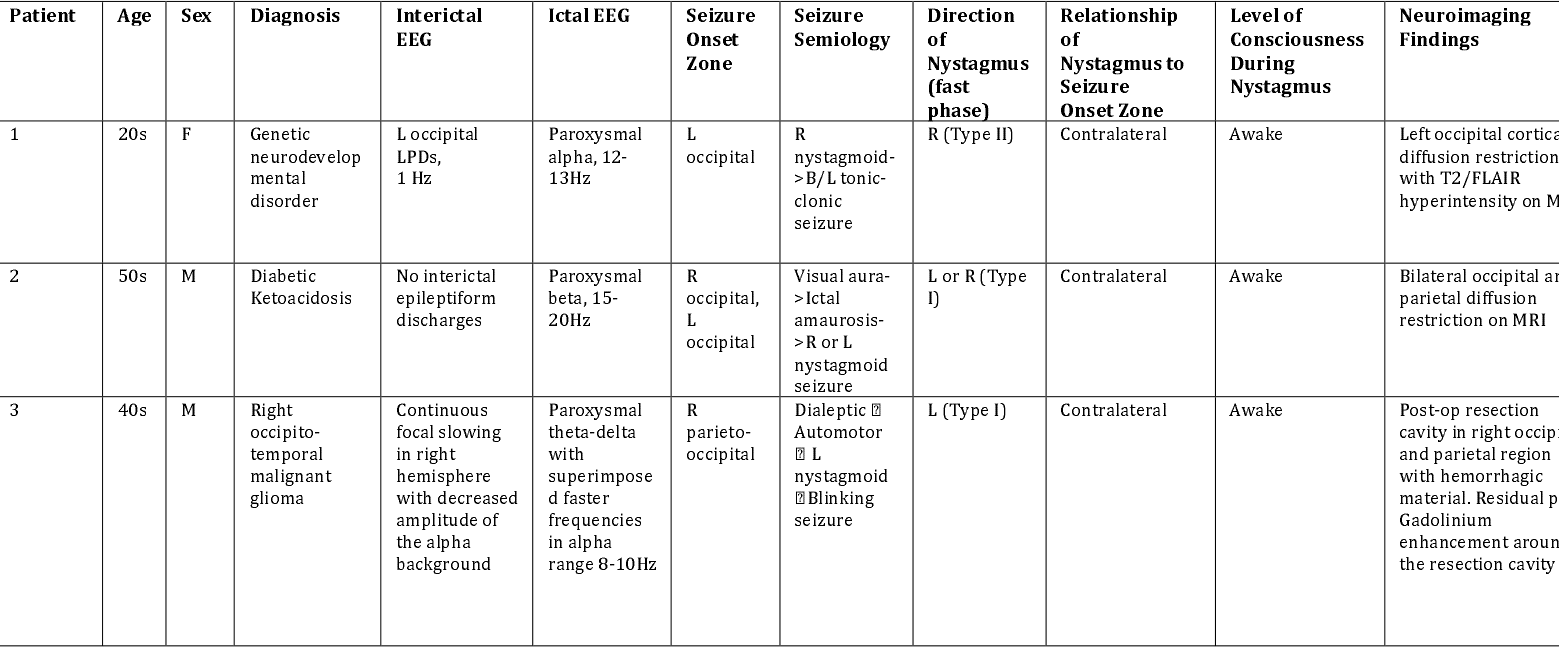
Patient demographics, characteristics of interictal & ictal EEG, seizure semiology, characteristics of nystagmus and relationship to seizure onset zone and neuroimaging F: female, M: male, L: left, LPD: lateralized periodic discharges, R: right

Patient #1’s EEG was consistent with a right beating nystagmus with leftward linear slow phases with left occipital seizure. With the prolonged TC, an overall initial deviation to the left was noted with rightward saccadic eye movements (Fig. 1a). Oculogram using F7-Cz demonstrated the eyes crossing the midline with both slow and fast phases (Fig. 1b). These characteristics are consistent with type II EN.

**Figure 1:**
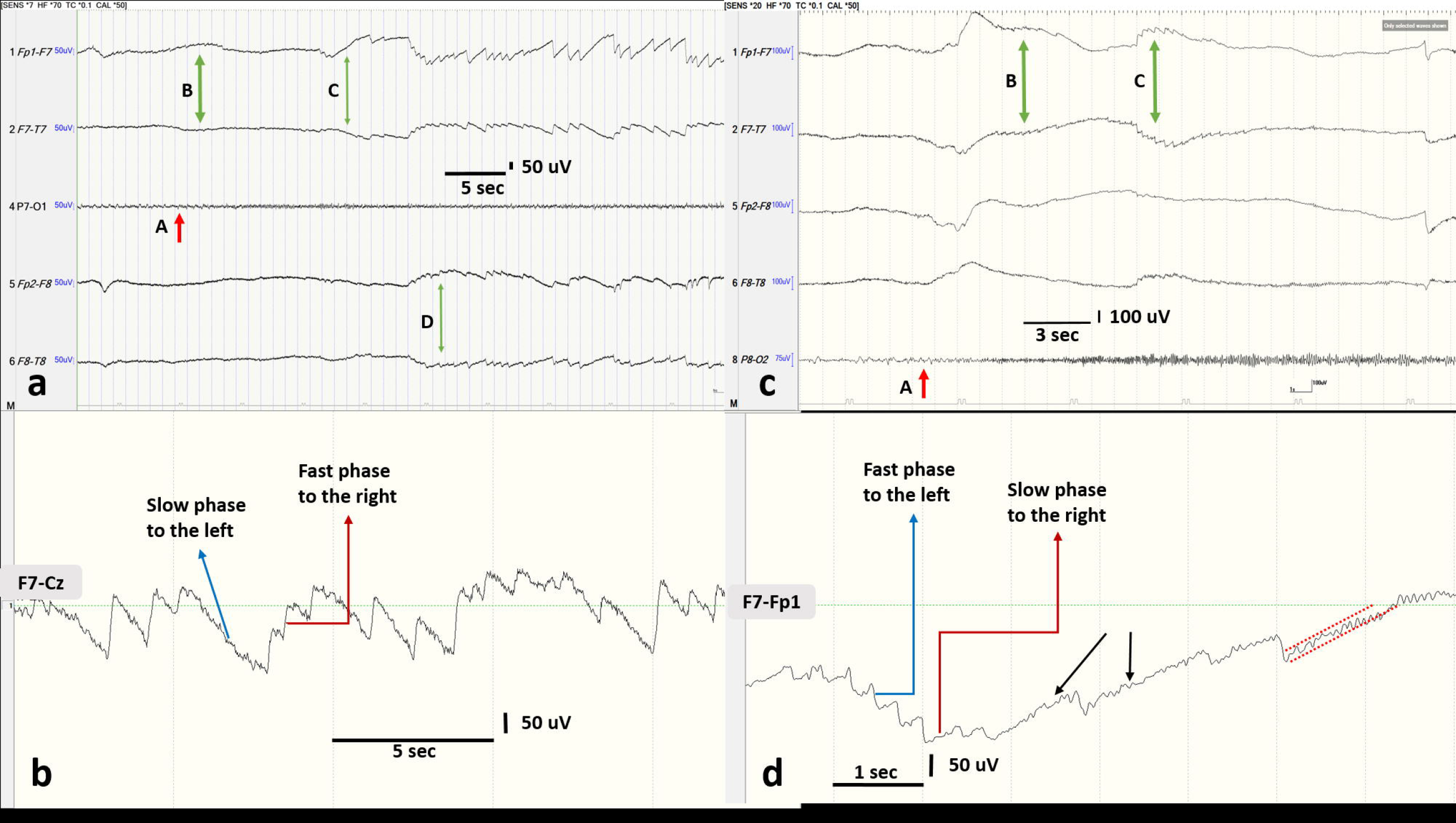
EEG analysis demonstrating epileptic nystagmus (EN). (a) EN in patient #1. 60s EEG page with left and right temporal channels demonstrating the nystagmus. Channel P7-O1 indicates the seizure onset with paroxysmal fast activity at A. The large positivity at F7 at points B & C indicates the initial leftward eye deviation. D indicates corrective rightward saccades. TC for the first two and last two channels is at 10s whereas that for P7-O1 is 0.1s. (b) Channel F7-Cz with TC of 10s demonstrating the oculogram for patient #1. Horizontal dotted green line represents the EEG baseline. Slow phases are mostly linear. Both slow and fast phases cross the midline. (c) EN in patient #2. 30s EEG page with left and right temporal channels demonstrating the nystagmus. Channel P8-O2 indicates the seizure onset with paroxysmal fast activity at A. The large positivity at F7 at points B & C indicates the overall leftward eye deviation. This deviation is produced by a ‘staircase’ of leftward saccades. TC for the first four channels is at 2s whereas that for P8-O2 is 0.1s. (d) Channel F7-Fp1 with TC of 2s demonstrating the oculogram for patient #2. Horizontal dotted green line represents the EEG baseline. The prolonged slow phases show a negative exponential decay approaching the baseline, as indicated by the two parallel dotted red lines. This is consistent with declining velocity as the eyes approach the midline. Black arrows indicate progressively longer slow phases.

In patients #2 and #3, the ictal EEG demonstrated repetitive saccadic eye movements contralateral to the seizure onset zone with overall eye deviation in the same direction as the saccades (Fig. 1c). Oculogram using F7-Fp1 demonstrated an overall contraversive deviation without crossing the midline (Fig. 1d). The slow phases were very brief, becoming progressively longer. The longer slow phases did show a waveform with a negative exponential slope consistent with decreasing slow phase velocity (Fig. 1d). These characteristics are consistent with type I EN.

The frequency of type II EN was 0.5-2Hz with a total duration of ∼2 minutes. The frequency of type I EN was 2-3Hz with a total duration of ∼15 seconds.

## 4. Discussion

Nystagmus is defined as abnormal repetitive oscillatory eye movements that interfere with vision (Leigh and Zee, 2015). Epilepsy is an uncommon cause of acquired nystagmus and EN is most commonly seen with seizures involving the posterior cortical regions (Kaplan and Tusa, 1993; Leigh and Zee, 2015). It is rarely associated with frontal lobe epilepsy (Lee et al.,2014). EN can be present in isolation as seizure semiology and has been reported at all ages (Nicita et al.,2010; Bekdik et al.,2006; Harris et al.,1997). In the majority of cases it is conjugate and horizontal (Kaplan and Tusa, 1993; Leigh and Zee, 2015).

As demonstrated by our results, type I EN is characterized by an overall deviation of the eyes contralateral to the seizure onset zone. A ‘staircase’ of contraversive saccades produces this deviation. As mentioned above, type I EN is postulated to be generated by activation of the cortical areas responsible for saccadic eye movements such as the frontal eye field (FEF) in posterior middle frontal gyrus or the parietal eye field (PEF) within the banks of the intraparietal sulcus (Kaplan and Tusa, 1993; Leigh and Zee, 2015). The activation of these areas is known to generate contraversive saccades with contraversive gaze deviation. A recent study has shown that in majority of versive seizures, the gaze deviation is saccadic rather than one smooth movement (Fotedar et al., 2022). The nystagmoid appearance is due to ipsiversive slow drifts alternating with the saccades, related to an additional gaze-holding defect, which accounts for the post-saccadic slow drift (Video 1). The velocity of the slow phase characteristically decreases as the eyes approach the midline and the eyes never cross the midline (Kaplan and Tusa, 1993; Tusa et al.,1990; Leigh and Zee, 2015).

Type II EN, on the other hand, consists of an initial ipsiversive eye deviation due to a slow movement, which is interrupted by corrective contraversive saccades (Video 2). The initial ipsiversive slow movement is produced by the activation of the cortical smooth pursuit region in the posterior superior temporal sulcus at the temporo-parieto-occipital (TPO) junction, known as middle temporal (MT) and medial superior temporal (MST) areas (Leigh and Zee, 2015). Experimental studies in monkeys have shown that activation of these areas results in an ipsiversive slow eye movement (Lüders and Noachtar, 2000). As demonstrated in the results, the slow phases are mostly linear and both fast and slow eye movements cross the midline (Kaplan and Tusa, 1993; Tusa et al., 1990; Leigh and Zee, 2015). We have summarized the characteristics of type I and II EN in Table 2.

**Table 2:** Comparison of major characteristics of Type I and Type II EN.

| **Type I EN** | **Type II EN** |
| --- | --- |
| Overall initial deviation contralateral to seizure onset zone | Overall initial deviation ipsilateral to seizure onset zone |
| The deviation is produced by a “staircase” of saccades | The deviation is produced by a slow movement |
| The eyes do not cross the midline | The eyes cross the midline |
| Prolonged slow phases have a negative exponential decay | Slow phases are mostly linear |
| Frequency is 2-3hz with a total duration of ~15 seconds | Frequency is 0.5-2Hz with a total duration of ~2 minutes |
| Likely mechanism is activation of saccade-generating areas like parietal eye field or frontal eye field | Likely mechanism is activation of the smooth pursuit region |

A third mechanism, similar to type II EN, is postulated to be related to the activation of the optokinetic region and can also produce a sensation of vertigo (Kaplan and Tusa, 1993; Tusa et al., 1990; García-Pastor et al., 2002; Ma et al., 2015). However, in all cases, the fast phase of the nystagmus is contralateral to the seizure onset zone (Lee et al., 2014; Nicita et al., 2010; Bekdik et al., 2006; Harris et al., 1997).

Previous studies suggest that, in awake patients, an ictal discharge frequency of ≥10Hz is associated with EN (Kaplan and Tusa, 1993). Our study is mostly in agreement with this observation.

## 5. Conclusions

In this study, we demonstrate a systematic method of eye movement analysis using EEG only that could help identify and classify EN in patients with occipital and parietal seizures without the help of additional EOG electrodes.

## Data Availability

All data produced in the present study are available upon reasonable request to the authors.

## Videos

**Video 1:** Eye movement animation representing type I EN. Video demonstrates leftward saccades alternating with rightward post-saccadic slow drifts without crossing the midline. This is representative of nystagmus seen in Figure 1c. The eye sketch is created using BioRender.com.

**Video 2:** Eye movement animation representing type II EN. Video demonstrates leftward slow phases alternating with rightward saccades crossing the midline. This is representation of nystagmus seen in Figure 1a. The eye sketch is created using BioRender.com.

## References

[1] Kaplan, P. W., & Tusa, R. J., 1993. Neurophysiologic and clinical correlations of epileptic nystagmus. Neurology, 43(12), 2508–2514. 10.1212/wnl.43.12.2508.

[2] Tusa, R. J., Kaplan, P. W., Hain, T. C., & Naidu, S., 1990. Ipsiversive eye deviation and epileptic nystagmus. Neurology, 40(4), 662–665. 10.1212/wnl.40.4.662.

[3] Lüders, H.O., Noachtar, S., 2000. Atlas and classification of electroencephalography. Saunders, Philadelphia.

[4] Leigh, R.J., Zee, D.S., 2015. The neurology of eye movements. Oxford University Press, New York.

[5] Lee, S. U., Suh, H. I., Choi, J. Y., Huh, K., Kim, H. J., & Kim, J. S., 2014. Epileptic nystagmus: A case report and systematic review. Epilepsy & behavior case reports, 2, 156–160. 10.1016/j.ebcr.2014.08.004

[6] Nicita, F., Papetti, L., Spalice, A., Ursitti, F., Massa, R., Properzi, E., & Iannetti, P., 2010. Epileptic nystagmus: description of a pediatric case with EEG correlation and SPECT findings. Journal of the neurological sciences, 298(1-2), 127–131. 10.1016/j.jns.2010.08.022.

[7] Bekdik, P., Sener, U., Asan, I. F., Ozcelik, M., & Zorlu, Y., 2006. Epileptic nystagmus. Epileptic disorders : international epilepsy journal with videotape, 8(4), 305–308..

[8] Harris, C. M., Boyd, S., Chong, K., Harkness, W., & Neville, B. G., 1997. Epileptic nystagmus in infancy. Journal of the neurological sciences, 151(1), 111–114. 10.1016/s0022-510x(97)00102-0.

[9] Fotedar, N., Gajera, P., Pyatka, N., Nasralla, S., Kubota, T., Vaca, G. F., Shaikh, A. G., & Lüders, H. O., 2022. A descriptive study of eye and head movements in versive seizures. Seizure, 98, 44–50. 10.1016/j.seizure.2022.04.003.

[10] García-Pastor, A., López-Esteban, P., & Peraita-Adrados, R., 2002. Epileptic nystagmus: a case study video-EEG correlation. Epileptic disorders : international epilepsy journal with videotape, 4(1), 23–28..

[11] Ma, Y., Wang, J., Li, D., & Lang, S., 2015. Two types of isolated epileptic nystagmus: case report. International journal of clinical and experimental medicine, 8(8), 13500–13507.

